# Rare-Variant Burden Analysis of Dystonia Genes in Parkinson’s Disease

**DOI:** 10.64898/2026.04.04.26349768

**Authors:** Sajanth Kanagasingam, Sitki Cem Parlar, Lang Liu, Ziv Gan-Or, Konstantin Senkevich

**Affiliations:** Department of Human Genetics, McGill University, Montréal, QC, Canada; The Neuro (Montreal Neurological Institute-Hospital), McGill University, Montréal, QC, Canada; Department of Neurology and Neurosurgery, McGill University, Montréal, QC, Canada; Department of Specialized Medicine, Division of Medical Genetics, McGill University Health Centre, Montreal, QC, Canada

## Abstract

**Background:** Dystonia frequently co-exists with Parkinson’s disease (PD), yet the extent of genetic overlap remains insufficiently explored.

**Objective:** To examine whether rare variants in dystonia-related genes are associated with PD or early-onset PD (EOPD).

**Methods:** We curated 44 dystonia-related genes using OMIM and the updated Movement Disorder Society report on hereditary dystonia. Whole-genome sequencing data from 5,315 PD patients, including 300 with EOPD, and 36,902 controls across the Accelerating Medicines Partnership–PD and UK Biobank cohorts were analyzed. For each gene, we evaluated rare variants (minor allele frequency <1%) in four pre-specified variant classes: exonic, nonsynonymous, CADD score ≥20 and loss-of-function. For the rare-variant burden analysis, SKAT-O was performed, followed by meta-analysis with MetaSKAT.

**Results:** In analyses of all PD cases, several genes showed nominal associations in meta-analysis: *SQSTM1* (*P*_loss-of-function_ = 5.52×10^−3^), *AOPEP* (*P*_exonic_ = 6.96×10^−3^; *P*_nonsynonymous_ = 0.017), *KCNA4* (*P*_exonic_ = 0.017), *SPR* (*P*_exonic_ = 0.029), *SLC30A10* (*P*_CADD ≥20_ = 0.046), and *ACTB* (*P*_exonic_ = 0.047). However, none remained significant after multiple-testing correction. In exploratory EOPD analyses, five genes reached significance after multiple test correction (*ATP5MC3, DNAJC12, KMT2B, TBC1D24, TMEM151A*). These signals were driven by small numbers of variants and were not robust to leave-one-variant-out analyses. *GCH1* was nominally significant in the meta-analysis of EOPD (*P*_nonsynonymous_ = 4.36×10^−3^, *P*_FDR_ = 0.062).

**Conclusions:** Rare variants in dystonia-related genes do not appear to make a major contribution to PD risk overall. Signals observed in the EOPD subset were based on small numbers of variant carriers and require replication in larger cohorts.

## Introduction

Dystonia is a complex, clinically and genetically heterogeneous movement disorder characterized by excessive and involuntary muscle contractions that produce abnormal movements or postures, with more than 250 genes potentially implicated.^1^ Even among carriers of the same genetic mutation, clinical expressivity is highly variable.^2^ Dystonia also frequently co-occurs with Parkinson’s disease (PD), with dystonic features reported in up to 30% of individuals with PD, either as an early manifestation or as a treatment-related complication.^3,4^ This overlap is particularly prominent in patients with early-onset PD (EOPD),^5^ and especially those with recessive monogenic forms such as *PARK7, PINK1,* and *PRKN*.^6^ In addition, PD patients with p.G2019S *LRRK2* develop dystonic features following levodopa initiation at a higher rate than patients with idiopathic PD.^7^ Conversely, certain dystonia syndromes, including dopa-responsive dystonia, can present with parkinsonism alongside childhood-onset dystonia, making it challenging to distinguish from EOPD.^8^

Both PD and dystonia involve dysfunction of nigrostriatal dopaminergic signaling within basal ganglia motor circuits.^9^ Several monogenic disorders present with combined dystonia-parkinsonism phenotypes, including those caused by variants in *GCH1*, *TH*, and *TAF1*.^10,11^ Notably, *GCH1* encodes a key enzyme in tetrahydrobiopterin synthesis, an essential cofactor for dopamine production, and pathogenic variants have been implicated in both dopa-responsive dystonia and PD as well as other movement disorders such as hereditary spastic paraplegia.^8,12–16^ Other genes associated with combined dystonia-parkinsonism syndromes, such as *ATP1A3*, have not been clearly linked to PD susceptibility.^17^

Several studies have reported enrichment of rare variants in dystonia-associated genes among PD cohorts.^12,18^ However, a large-scale gene-level burden analysis of dystonia-related genes across European PD cohorts has not yet been performed. While genome-wide association studies have identified 134 risk loci for PD,^19^ comparable studies in dystonia have not detected robust signals, suggesting that dystonia may be driven predominantly by rare genetic variation.^20^ Accordingly, the contribution of dystonia-related genes to PD is best evaluated using rare-variant approaches. In this study, we systematically assess whether genes implicated in dystonia are enriched for rare variation in large PD cohorts.

## Methods

### Population

The study population comprised of 5,315 PD patients and 36,902 controls from two cohorts, the Accelerating Medicines Partnership–Parkinson Disease (AMP-PD) initiative cohorts and the UK Biobank (UKBB) cohort. Only participants of European ancestry were included in both cohorts given the limited representation of other ancestry groups. Any first and second-degree relatives were excluded from the analysis. In the UKBB cohort, individuals were included based on their self-report data (field 20002), hospital inpatient ICD10 diagnoses (field 41270) and, when present, primary and secondary causes of death from their death registry (fields 40001, 40002). PD proxies, identified via available data for illnesses of father, mother and siblings (fields 20107, 20110, 20111), were excluded from controls.

A total of 300 early-onset PD cases (EOPD, ≤50 years) were also identified, based on the algorithmically defined date of PD report (field 42032) in the UKBB cohort and age at baseline evaluation in the AMP-PD cohort. Demographic details on these cohorts can be found in Table 1.

**Table 1.**
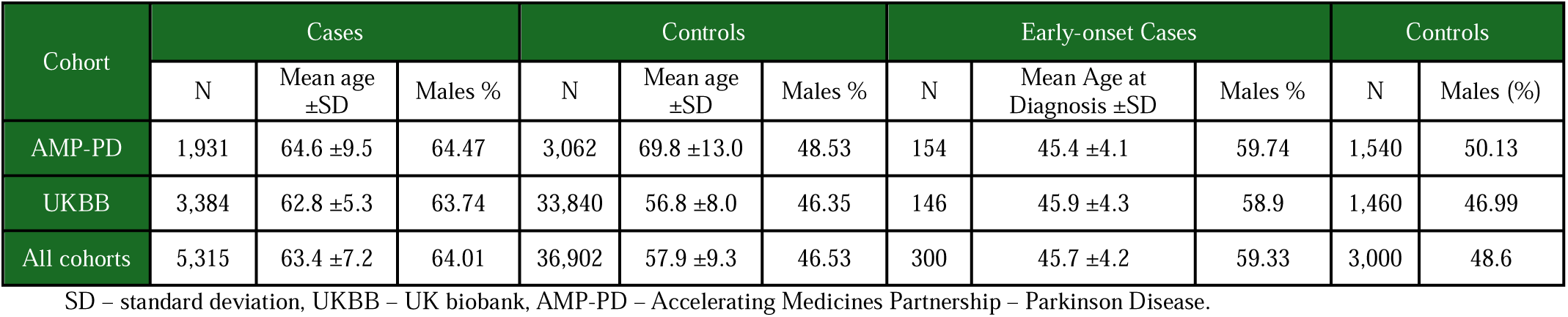
Demographics.

McGill University’s *Faculty of Medicine Institutional Review Board A* approved the study protocol (REB file number 21-11-023). This study utilized de-identified data; primary informed consent was obtained from all participants by the respective contributing sites.

### Gene Selection

To generate a comprehensive list of dystonia-related genes, the Online Mendelian Inheritance in Man (OMIM) compendium and the recommendations of the International Parkinson and Movement Disorder Society (MDS) Task Force on the Nomenclature of Genetic Movement Disorders were consulted. Gene entries were extracted from OMIM Gene Map using the search term “dystonia”, from the OMIM dystonia phenotypic series (PS128100), and from the MDS updated list of hereditary dystonia (isolated and combined, excluding complex).^21^ Dystonia loci that do not refer to a specific identified gene in the HUGO nomenclature and X-linked genes were removed as they would require separate statistical analyses. This process yielded the following 44 dystonia-related genes: *ACTB, ANO3, AOPEP, ATP1A3, ATP5MC3, COL6A3, COX20, DNAJC12, EIF2AK2, GCH1, GNAL, GNAO1, HPCA, KCNA4, KCNN2, KCTD17, KMT2B, MECR, NR4A2, NUP54, PLEKHG2, PNKD, PRKRA, PRRT2, SCP2, SGCE, SHQ1, SLC18A2, SLC2A1, SLC30A10, SLC39A14, SLC6A3, SPR, SQSTM1, TBC1D24, TH, THAP1, TMEM151A, TOR1A, TSPOAP1, TUBB4A, VPS11, VPS16, WARS2*.

### Whole-genome sequencing data quality control and annotation

The genetic data in AMP-PD and UKBB were aligned to the human reference genome hg38, and canonical GENCODEV48 coordinates for each of the 44 dystonia-related genes were obtained via UCSC’s Table Browser.^22^ In addition to the quality control performed by the source institutions,^23,24^ standard quality control procedures were applied using plink 2.0 and bcftools 1.16. Briefly, variants with genotype quality <25, read depth <25, or missingness >5% were filtered out. An exception applies to the AMP-PD data, where only read depth and missingness filters were applied as genotype quality information was not available. In both cohorts, rare variants were then filtered using a minor allele frequency (MAF) cut-off of 0.01.

The filtered variants were annotated using Ensembl Variant Effect Predictor (VEP)^25^ v.114 to obtain their AlphaMissense pathogenicity scores,^26^ Combined Annotation Dependent Depletion (CADD)^27^ v.1.7 Phred scores, and ClinVar clinical significance.^28^ AlphaMissense scores were used in single-variant analysis to identify pathogenic variants by applying a threshold of 0.564. This cutoff corresponds to an estimated 90% precision based on ClinVar annotations.^26^

### Rare-Variant Analysis

To analyze the rare variants, the optimized sequence Kernel association test (SKAT-O, R package) was applied in each cohort separately, followed by meta-analysis with the MetaSKAT R package.^29,30^ A sub-analysis of EOPD cases was conducted using the same approach. All analyses were adjusted for age, sex, and top 5 principal components. To reduce type I error from large case-control imbalances in the UKBB cohort and the EOPD subanalyses, the case-control ratio was fixed at 1:10 by randomly sampling controls. However, to provide additional context at the individual variant level, single-variant frequency estimates (MAF and odds ratios) for nominated variant candidates were calculated using the full UK Biobank cohort (3,384 cases and 338,955 controls) rather than the down-sampled dataset.

Burden analyses were performed for each of the 44 dystonia genes individually, followed by a global analysis of all 44 genes together. For each analysis, four variant sets were defined. a) Exonic variants. b) Nonsynonymous variants. c) CADD: variants with a CADD Phred score greater or equal to 20. d) Loss-of-function variants with a high impact as defined by VEP, including frameshift, stop gain, stop-loss, start-loss, splice donor/acceptor, and transcript ablation. False discovery rate correction (FDR) using the Benjamini–Hochberg method was then applied to the aggregate of all p-values generated within each cohort. Corrected *P*_FDR_ < 0.05 was considered significant, while uncorrected *P* < 0.05 was reported as nominal. It is important to note that variant sets containing less than two variants were excluded from the analysis, as such cases are not compatible with SKAT-based aggregation frameworks.

### Variant Nomination

To investigate the variants driving the SKAT-O associations, a shortlist for leave-one-out sensitivity analysis was generated by comparing individual variant frequencies between cases and controls. Odd ratios (OR) were also calculated for this purpose, both with and without Yates’ continuity correction (adding a constant of 0.5 to all values). The leave-one-out analysis was then conducted by sequentially excluding individual variants until their respective variant sets no longer achieved significance.

## Results

### Rare variants in dystonia-related genes are not associated with PD overall

For the rare-variant burden analysis, we identified 157,086 rare variants across both cohorts, including 4,142 nonsynonymous and 374 loss-of-function variants (Supplementary Table 1). In AMP-PD, nominal associations were detected for *KCNA4* (*P*_exonic_ = 3.84×10^−3^)*, SQSTM1* (*P*_CADD_ = 5.76×10^−3^; *P*_nonsynonymous_ = 0.021), and *THAP1* (*P*_exonic_ = 6.43×10^−3^). In UKBB, nominal signals were observed for *AOPEP* (*P*_exonic_ = 2.42×10^−3^; *P*_nonsynonymous_ = 0.010) and *COX20* (*P*_nonsynonymous_ = 0.020). None of these associations survived FDR correction (Supplementary Table 2). The global burden analysis of all 44 dystonia genes also did not reveal any significant association (Supplementary Table 3).

Cross-cohort meta-analysis using MetaSKAT identified several gene sets reaching nominal significance, including *SQSTM1* (*P*_loss-of-function_ = 5.52×10^−3^), *AOPEP* (*P*_exonic_ = 6.96×10^−3^; *P*_nonsynonymous_ = 0.017), *KCNA4* (*P*_exonic_ = 0.017), *SPR* (*P*_exonic_ = 0.029), *SLC30A10* (*P*_CADD_ = 0.046), and *ACTB* (*P*_exonic_ = 0.047). However, none remained significant after FDR correction.

Leave-one-out sensitivity analysis demonstrated that the two strongest signals, *SQSTM1* and *AOPEP*, were each driven by a single variant: p.Gln310Ter and p.Arg594Trp, respectively. The *SQSTM1* p.Gln310Ter variant was only observed in the UKBB cohort (MAF cases = 1.49×10^−4^, MAF controls = 0). The *AOPEP* p.Arg594Trp variant was present in both AMP-PD (MAF cases = 2.59×10^−4^, MAF controls = 1.63×10^−4^; OR 1.59, 95% CI 0.16–15.26) and UKBB (MAF cases = 1.37×10^−3^, MAF controls = 3.78×10^−4^; OR 3.64, 95% CI 1.87–7.09). In UKBB, it exceeded the reported frequency in non-Finnish Europeans according to gnomAD (3.97×10^−4^). The clinical relevance of p.Arg594Trp remains uncertain, as in silico predictions were inconsistent (CADD Phred = 26 and low AlphaMissense score).

Additional single-variant analysis evaluated variants previously classified as pathogenic by ClinVar and the American College of Medical Genetics and Genomics (ACMG)^31^ criteria within the GP2 dataset.^12^ *GCH1* p.Lys224Arg was observed in both AMP-PD (MAF cases = 7.77×10^−4^, MAF controls = 4.90×10^−4^; OR 1.59, 95% CI 0.32-7.87) and UKBB (MAF cases = 6.19×10^−4^, MAF controls = 3.48×10^−4^; OR 1.78, 95% CI 0.66–4.79). *GCH1* p.Val204Ile was detected in UKBB (MAF cases = 1.54×10^−4^, MAF controls = 1.53×10^−4^; OR 1.01, 95% CI 0.14–7.26). *TOR1A* p.Glu303del was identified in one case from AMP-PD (MAF cases = 2.59×10^−4^) and in UKBB controls (MAF controls = 1.94×10^−5^). The remaining previously nominated pathogenic variants were identified exclusively in UKBB controls: *GCH1* p.Arg249Ser (MAF 1.54×10^−6^) and p.Arg235Trp (1.54×10^−6^); *THAP1* p.Lys73Thr (1.22×10^−5^); *TOR1A* p.Arg288Gln (1.11×10^−4^); as well as *VPS16* p.Gln127Ter (1.52×10^−6^) and c.2375+1G>A (1.85×10^−5^).

### Gene-level signals identified in EOPD analyses

In the EOPD subanalysis, we identified 28,808 rare variants across both cohorts, including 738 nonsynonymous variants and 49 loss-of-function variants (Supplementary Table 4). Rare-variant burden testing was once again performed using SKAT-O within each cohort, followed by cross-cohort meta-analysis (Supplementary Table 5). Five genes met the FDR-adjusted significance threshold in specific variant sets: *TBC1D24* (*P*_loss-of-function_ = 6.61×10^−5^, *P*_FDR_ = 8.47×10^−3^), *DNAJC12* (*P*_CADD_ = 4.31E×10^−4^, *P*_FDR_ = 0.027), *KMT2B* (*P*_CADD_ = 7.12×10^−4^, *P*_FDR_ = 0.027), *ATP5MC3* (*P*_nonsynonymous_ = 9.69×10^−4^, *P*_FDR_ = 0.027), and *TMEM151A* (*P*_CADD_ = 1.17×10^−3^, *P*_FDR_ = 0.027; *P*_nonsynonymous_ = 1.24×10^−3^, *P*_FDR_ = 0.027). While several of these sets showed nominal signals in individual cohorts, they only surpassed the threshold for corrected significance when combined in meta-analysis (Table 2).

**Table 2.**
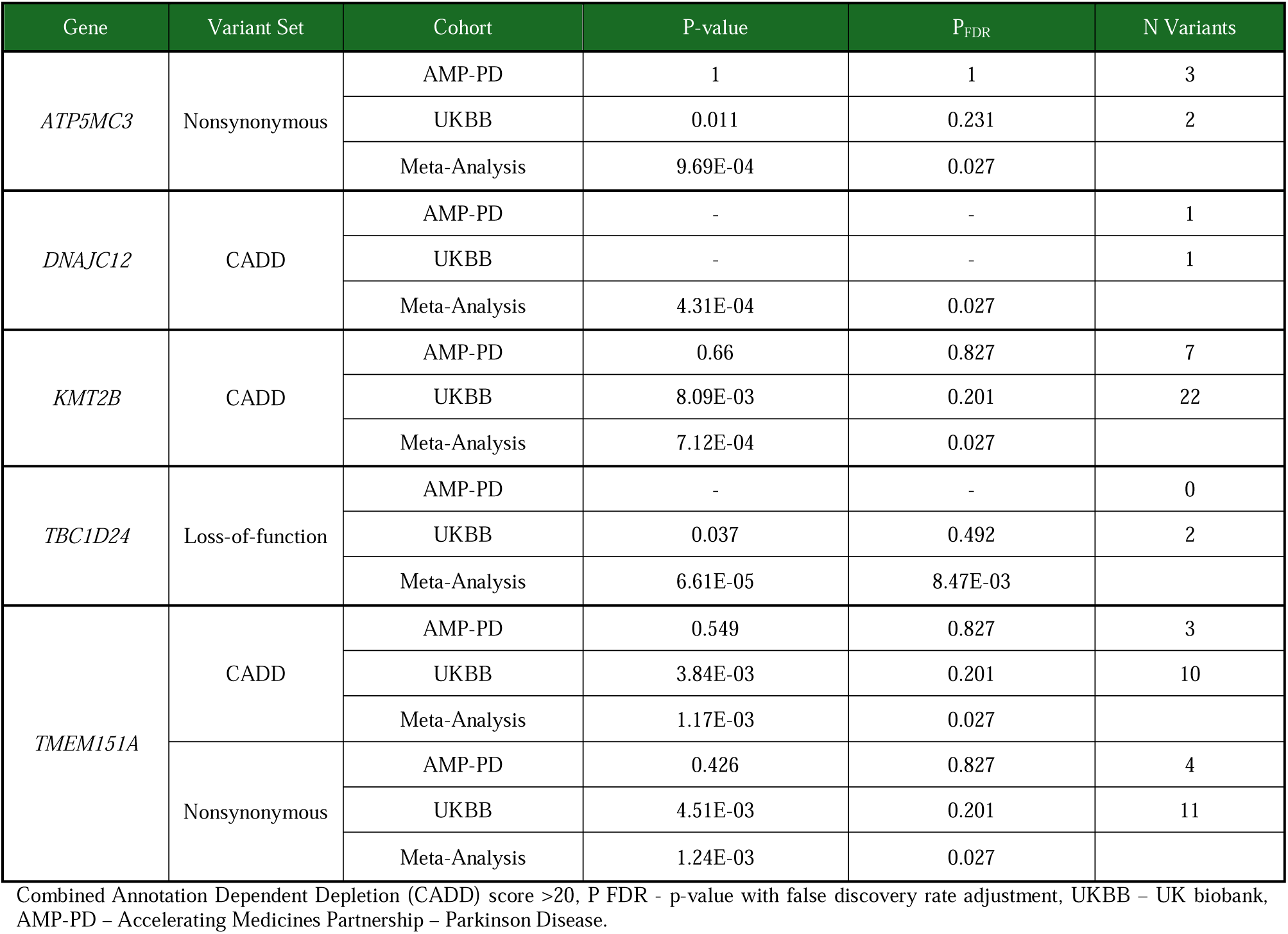
Significant genes amongst EOPD cases in meta-analysis with corresponding cohort-level findings.

Leave-one-out sensitivity analysis indicated that these associations were highly sensitive to single-variant removal (Supplementary Table 6), consistent with the limited number of EOPD carriers per variant set. Specifically, signals for *TBC1D24, DNAJC12,* and *KMT2B* were each driven by a single variant (c.-115-2A>G, p.Arg74Cys, and p.Pro2293Ser, respectively), whereas two variants (p.Ile65Val and p.Lys6Asn) accounted for the signal observed in *ATP5MC3*. The *TMEM151A* signal reflected contributions from several variants (p.Arg277Cys, p.Ala77Thr, and p.Arg187His). Evaluation of variant frequencies in the full UKBB dataset (Supplementary Table 7) and comparison with gnomAD did not support enrichment of these variants within PD. Consistently, the global burden analysis revealed no significant associations, although a nominal signal was identified for the high-CADD variant set in the meta-analysis (*P*_CADD_ = 0.023, *P*_FDR_ = 0.069) (Supplementary Table 8).

Additionally, it is noteworthy that *GCH1* was nominally significant at the cohort-level (AMP-PD: *P*_nonsynonymous_ = 0.032; UKBB: *P*_CADD_ = 0.017) but did not survive FDR correction in meta-analysis (*P*_nonsynonymous_ = 4.36×10^−3^, *P*_FDR_ = 0.062; *P*_CADD_ = 9.18×10^−3^, *P*_FDR_ = 0.118).

## Discussion

In this study, we systematically examined the contribution of rare variants in dystonia-related genes to PD and EOPD. This large-scale rare-variant burden analysis of 44 dystonia genes across 5,315 PD cases and 36,902 controls of European ancestry did not identify significant enrichment of rare variants in dystonia genes after multiple testing correction. However, several genes demonstrated consistent nominal signals in meta-analysis, including *SQSTM1* and *AOPEP*, for which sensitivity analyses identified specific variants driving their association with PD. These findings are consistent with recent work by GP2, which reported pathogenic variants in dystonia genes in only 0.54% of PD patients, with *GCH1* being the most frequently implicated gene.^12^ Similarly, burden analysis in a Chinese PD cohort identified only suggestive, non-significant associations between dystonia genes and PD.^18^ Together, these data suggest that truly pathogenic dystonia variants are rare in unselected PD populations and that the genetic overlap between dystonia and PD is limited at the gene-burden level despite clinical and pathway-level convergence.

Among dystonia genes, *GCH1* stands out as the most consistently validated contributor to PD risk.^12–15^ In the present study, previously reported pathogenic *GCH1* variants (p.Lys224Arg, p.Val204Ile) were observed at low frequencies in both cases and controls without significant enrichment, consistent with their status as rare risk factors rather than highly penetrant causal variants. The nominal *GCH1* signal observed in EOPD, while not surviving FDR correction, is consistent with its established role as a rare contributor to both dystonia and parkinsonism phenotypes. This near-significant result may potentially reflect insufficient statistical power in the EOPD analysis, underscoring the need for larger cohorts in future studies.

In the EOPD subanalysis, five genes (*TBC1D24, DNAJC12, KMT2B, ATP5MC3,* and *TMEM151A*) achieved FDR-corrected significance in specific variant sets. These genes are established causes of dystonia or combined movement disorder syndromes but have not been consistently implicated in isolated PD.^32–36^ However, these associations should be interpreted with considerable caution for several reasons. First, leave-one-out analyses demonstrated that most signals were driven by single variants, raising concerns about robustness. Second, comparison with gnomAD population frequencies did not support enrichment of these variants in PD. Third, the small sample size limits statistical power and increases susceptibility to false-positive findings.

*SQSTM1/p62* is a multifunctional autophagy adaptor protein implicated in multiple neurodegenerative diseases, including PD, through its roles in autophagy and protein aggregate clearance.^37^ The relevance of *SQSTM1* for PD pathology is reinforced by its interactions with *LRRK2*, a key player in both familial and sporadic forms of PD.^38^ Pathogenic variants in *LRRK2* have been shown to increase *SQSTM1* phosphorylation at Thr138 and, conversely, *LRRK2* autophagic degradation is increased in cell models where *SQSTM1* is overexpressed.^39,40^ However, pathogenic *SQSTM1* variants were previously associated with amyotrophic lateral sclerosis, frontotemporal dementia, distal myopathy, and Paget’s disease of bone but not with PD.^41^ Replication in independent cohorts would be required to clarify its relevance.

*AOPEP* encodes aminopeptidase O, a zinc-requiring proteolytic enzyme highly expressed in neuroglia, and is a recently identified cause of autosomal recessive dystonia (Zech-Boesch syndrome).^42^ While the clinical literature predominantly describes biallelic loss-of-function mutations leading to isolated multifocal or generalized dystonic syndromes, a minority of patients exhibit a combined dystonia-parkinsonism phenotype.^43,44^ Indeed, three individuals of French and Chinese ancestries have been reported to present with akinesia and freezing of gait in addition to their generalized dystonia. However, all three of these patients were found to have biallelic nonsense mutations as opposed to the missense variant of unknown significance we identified in this study. Therefore, independent evaluation of variants in this gene in PD would be required to confirm this finding.

In terms of limitations, our study design necessitates caution in several areas. First, our analysis was restricted to autosomal genes, explicitly excluding X-linked loci such as *TAF1* as they would require separate statistical analysis because of differences in allele dosage. Consequently, significant sex-linked genetic contributors to the dystonia-PD genetic overlap may have been missed. Second, samples were restricted to European ancestry which limits the generalizability of our findings to other populations and precludes the identification of ancestry-specific PD risk variants. Third, slight differences in sequencing platforms, pre-processing quality control pipelines, and covariate definitions between the AMP-PD and UKBB cohorts may have introduced bias into our meta-analysis. Finally, reiterating caution for the EOPD subanalysis findings, these gene-level signals were from a small cross-cohort population of 300 EOPD cases and were driven by sparse variant counts, as shown by their sensitivity to leave-one-out analysis. We therefore consider this analysis exploratory, and replication in larger and more deeply phenotyped EOPD cohorts would be warranted to establish their validity.

In conclusion, our findings indicate that rare variants in dystonia-related genes are not major contributors to PD risk. Although several nominal signals were observed, they were driven by individual variants and require independent replication. These findings suggest that pathogenic variants in established dystonia genes are uncommon in PD cohorts.

## Funding agencies

This study was supported by grants from the Galen and Hilary Weston Foundation, the Michael J. Fox Foundation, the Canadian Consortium on Neurodegeneration in Aging (CCNA), the Canada First Research Excellence Fund (CFREF). The study received contributions from the G-Can (GBA1-Canada) Initiative. G-Can is supported by The Hilary and Galen Weston Foundation, Silverstein Foundation, and J. Sebastian van Berkom and Ghislaine Saucier. S.K. is supported by the Medical Class of 1989 Research Bursary from McGill University’s Faculty of Medicine and Health Sciences. Z.G.-O. holds a Fonds de recherche du Québec - Santé (FRQS) Chercheurs-boursiers award and is a William Dawson Scholar.

## Acknowledgment

We thank the patients and their families for participating in this study. S.K. is supported by the Medical Class of 1989 Research Bursary from McGill University’s Faculty of Medicine and Health Sciences. Z.G.O. is supported by the Fonds de recherche du Québec—Santé (FRQS) Chercheurs-boursiers award, in collaboration with Parkinson Quebec, and is a William Dawson Scholar.

Data used in the preparation of this article were obtained from the Accelerating Medicine Partnership® (AMP®) Parkinson’s Disease (AMP PD) Knowledge Platform. Data for this article were also obtained in January 2023 from the AMP-PD Knowledge Platform as per release 2.5. Since this release, more recent versions have been made available. The AMP® PD program is a public-private partnership managed by the Foundation for the National Institutes of Health and funded by the National Institute of Neurological Disorders and Stroke (NINDS) in partnership with the Aligning Science Across Parkinson’s (ASAP) initiative; Celgene Corporation, a subsidiary of Bristol-Myers Squibb Company; GlaxoSmithKline plc (GSK); The Michael J. Fox Foundation for Parkinson’s Research; Pfizer Inc.; AbbVie Inc.; Sanofi US Services Inc.; and Verily Life Sciences. ACCELERATING MEDICINES PARTNERSHIP and AMP are registered service marks of the U.S. Department of Health and Human Services.

Data used in the preparation of this article were obtained from the Accelerating Medicine Partnership® (AMP®) Parkinson’s Disease (AMP PD) Knowledge Platform. The AMP® PD program is a public-private partnership managed by the Foundation for the National Institutes of Health and funded by the National Institute of Neurological Disorders and Stroke (NINDS) in partnership with the Aligning Science Across Parkinson’s (ASAP) initiative; Celgene Corporation, a subsidiary of Bristol-Myers Squibb Company; GlaxoSmithKline plc (GSK); The Michael J. Fox Foundation for Parkinson’s Research; Pfizer Inc.; AbbVie Inc.; Sanofi US Services Inc.; and Verily Life Sciences. ACCELERATING MEDICINES PARTNERSHIP and AMP are registered service marks of the U.S. Department of Health and Human Services. Clinical data and biosamples used in preparation of this article were obtained from the (i) Michael J. Fox Foundation for Parkinson’s Research (MJFF) and National Institutes of Neurological Disorders and Stroke (NINDS) BioFIND study, (ii) Harvard Biomarkers Study (HBS) and the Stephen & Denise Adams Center for Parkinson’s Disease Research of Yale School of Medicine (CPDR-Y), (iii) National Institute on Aging (NIA) International Lewy Body Dementia Genetics Consortium Genome Sequencing in Lewy Body Dementia Case-control Cohort (LBD), (iv) NINDS Parkinson’s Disease Biomarkers Program (PDBP), and (v) MJFF Parkinson’s Progression Markers Initiative (PPMI). BioFIND is sponsored by The Michael J. Fox Foundation for Parkinson’s Research (MJFF) with support from the National Institute for Neurological Disorders and Stroke (NINDS). The BioFIND Investigators have not participated in reviewing the data analysis or content of the manuscript. Genome sequence data for the Lewy body dementia case-control cohort were generated at the Intramural Research Program of the U.S. National Institutes of Health. The study was supported in part by the National Institute on Aging (program #: 1ZIAAG000935) and the National Institute of Neurological Disorders and Stroke (program #: 1ZIANS003154). The Harvard Biomarker Study (HBS) is a collaboration of HBS investigators and funded through philanthropy and NIH and Non-NIH funding sources. The Stephen & Denise Adams Center for Parkinson’s Disease Research of Yale School of Medicine is funded through philanthropy and NIH and non-NIH funding sources. The HBS and CPDR-Y Investigators have not participated in reviewing the data analysis or content of the manuscript. PPMI is sponsored by The Michael J. Fox Foundation for Parkinson’s Research and supported by a consortium of scientific partners. The PPMI investigators have not participated in reviewing the data analysis or content of the manuscript. The Parkinson’s Disease Biomarker Program (PDBP) consortium is supported by the National Institute of Neurological Disorders and Stroke (NINDS) at the National Institutes of Health. The PDBP investigators have not participated in reviewing the data analysis or content of the manuscript.

This research used the NeuroHub infrastructure and was undertaken thanks in part to funding from the Canada First Research Excellence Fund, awarded through the Healthy Brains, Healthy Lives initiative at McGill University. And our sincere thanks to the participants and researchers of the UK Biobank, who made this effort possible. This project was completed under UK Biobank application 45551. This research was enabled in part by support provided by Calcul Québec and the Digital Research Alliance of Canada.

## Authors’ Roles

(1) Research project: A. Conception, B. Organization, C. Execution; (2) Statistical Analysis: A. Design, B. Execution, C. Review and Critique; (3) Manuscript Preparation: A. Writing of the first draft, B. Review and Critique.

S.K.: 1A, 1B, 1C, 2A, 2B, 3A.

S.C.P.: 1C, 2C, 3B

L.L.: 1C, 3B

Z.G.-O.: 1C, 2C, 3B.

K.S.: 1A, 1B, 2C, 3B.

## Financial Disclosures of all authors (for the preceding 12 months)

Z.G.-O. received consultancy fees from Lysosomal Therapeutics Inc., Idorsia, Prevail Therapeutics, Inceptions Sciences (now Ventus), Neuron23, Handl Therapeutics, UCB, Capsida, Vanqua Bio, Congruence Therapeutics, Ono Therapeutics, Denali, Bial Biotech, Bial, EG427, Takeda, Jazz Pharmaceuticals, Guidepoint, Lighthouse, and Deerfield. K.S. received consultancy fees from Acurex. None of these companies were involved in any parts of preparing, drafting, and publishing this study. The other authors have nothing to disclose.

## Data Availability

All supplementary materials, including additional tables and figures, are available in the Supporting Information section. The scripts used to generate the results from this study have been published in a public repository (https://github.com/Sajanth1/Rare-Variant-Analysis). The AMP-PD data was assessed using the Terra platform (https://amp-pd.org/). The UKBB data was acquired using WGS data from the UK Biobank Research Analysis Platform (https://www.ukbiobank.ac.uk/) under application number 45551.

